# Stroke in adults with primary intracranial tumours

**DOI:** 10.1101/2024.10.06.24314978

**Authors:** Stuart C. Innes, Lucia Yin, Gerald T. Finnerty

## Abstract

**BACKGROUND:** Malignant primary intracranial tumours are associated with stroke. Knowing how stroke differs in patients with either benign or malignant primary intracranial tumours would give insights into how intracranial tumours affect stroke and would help to improve the limited stroke guidelines for these patients.

**METHODS:** We conducted a retrospective cohort study of patients with benign or malignant primary intracranial tumours admitted with stroke (2011 - 2022) to a single centre with regional stroke and neuro-oncology units. Data collected included: stroke aetiology, stroke timing relative to tumour diagnosis, pre-stroke disability, post-stroke disability, stroke recurrence and treatment.

**RESULTS:** We identified 258 patients who had an index stroke (120 haemorrhagic, 135 ischaemic, three coincident haemorrhagic/ischaemic) at or after the diagnosis of their primary intracranial tumour (71% benign, 29% malignant). Symptomatic intracranial haemorrhage was more commonly associated with malignant primary intracranial tumours. Patients with a malignant primary intracranial tumour were 2.7 times more likely to have a stroke than patients with a benign intracranial tumour. We estimated that the risk of ischaemic stroke and symptomatic intracranial haemorrhage within six months of diagnosis of a benign primary intracranial were respectively 2.5 and 5.0 times that of the general population. In-hospital mortality and level of disability at hospital discharge (median modified Rankin scale score, 4) were similar for patients with benign or malignant primary intracranial tumours. Stroke recurrence was 22% at one year. Statins were associated with reduced stroke recurrence.

Patients with malignant tumours and high post-stroke disability were less likely to receive chemotherapy after hospital discharge.

**CONCLUSIONS:** Our findings suggest that stroke is a wider issue for patients with primary intracranial tumours than commonly recognised. Our data indicated that statins may help to prevent stroke recurrence. Despite concerns about intracranial haemorrhage, antiplatelet agents should be considered after ischaemic stroke.

---

Primary intracranial tumours are a heterogenous group of tumours affecting the brain, cranial nerves and cranial meninges. Common symptoms at clinical presentation include: seizures, headache, cognitive impairment and focal neurological disturbance.^1^ The current standard of care for intracranial tumours involves a combination of surgery, radiotherapy and chemotherapy.^2,3^ Much effort has been made to improve tumour treatment. Less attention has been paid to the neurological complications of these tumours and their treatment, such as stroke.^4^ This is surprising because patients with cancer have a greater risk of stroke than the general population.^5-9^

When different cancers are compared, patients with cancers of the central nervous system are particularly likely to have an ischaemic or haemorrhagic stroke that results in admission^8^ and are more likely to die within one year of their stroke.^9^ As a result, stroke in patients with malignant primary intracranial tumours represents a major management challenge.

Far less is known about stroke in patients with benign primary intracranial tumours despite benign intracranial tumours being six to seven times more prevalent than malignant primary intracranial tumours in adults.^10^ This is unexpected because a comparison of stroke in malignant and benign primary brain tumours could give insights into how tumour malignancy, tumour treatment and primary intracranial tumours in general affect stroke. Moreover, this knowledge would inform tailored management of stroke in patients with primary intracranial tumours.

Guidelines for primary and secondary stroke prevention in patients with primary intracranial tumours requires knowledge of: the type and timing of strokes, the consequences of stroke for patients and stroke recurrence.

Valuable information has come from studies in specialist neuro-oncology centres, which have reported clinical presentation, mortality and level of disability following ischaemic and haemorrhagic stroke.^7,11-14^ However, these studies have not reported separate results for malignant and benign primary intracranial tumours. Furthermore, the patient cohorts may have been biased towards patients who benefited from more therapeutic interventions and may not include all patients with stroke if the centre does not have a regional stroke unit. Data from cancer registries, which reflect the general population better, have suggested that patients with central nervous system malignancies have a worse prognosis after stroke,^8,9^ but do not provide the detailed information required for stroke guidelines.

Studies based on stroke registries have revealed important information on occult cancer in stroke patients and have suggested that ischaemic and haemorrhagic strokes are more common in patients with malignant intracranial tumours^15^ and are more likely to present with a stroke involving multiple vascular territories.^16^ More recently, a study combining regional stroke and cancer registries suggested that patients with cancer and an ischaemic stroke have a worse prognosis.^17^

The consequence of the lack of necessary data is that the guidelines for prevention and management of stroke in patients with primary intracranial tumours are limited.^18-21^

We investigated whether stroke profiles, including type and timing, differed between patients with a malignant or benign primary intracranial tumour. Our data indicate that tumour malignancy affects stroke type. However, stroke outcomes and stroke recurrence were similar. Our findings suggest points where stroke care may be improved in patients with primary intracranial tumours.

## METHODS

Adult (≥ 18 years old) patient data were obtained from a hospital with both a Regional Neuro-oncology service and Regional Stroke service by searching for patients with a primary intracranial tumour who had an inpatient stay between December 2011 to April 2022 due to a spontaneous or post-operative stroke affecting the brain.

A primary intracranial tumour was defined as a primary tumour of the brain, cranial nerves or cranial meninges using the 2021 World Health Organisation (WHO) Classification of Tumours of the Central Nervous System.^22^ Tumour diagnoses before 2021 were updated to the 2021 classification where possible. Brain tumours were divided into malignant (WHO grades 3, 4) and benign (WHO grades 1, 2) tumours. Some tumours showed evidence of malignant progression during the study. The tumour was classified as malignant at stroke diagnosis if the tumour was: 1) classified histopathologically as WHO grade 3 or above; 2) showed evidence of malignant progression on neuroimaging; or 3) was treated clinically for malignant progression. The ICD-10 codes for primary tumours of the brain, cranial nerves or cranial meninges were: C70.0, C70.9, C71.x, C72.2-5, C72.9, C75.1-3, C85.7, D32.0, D32.9, D33.0-3, D.33.7, D33.9, D35.2-4, D43.0-3, D43.7, D43.9, D44.4.

The index stroke was defined as the first clinical presentation of a stroke at or after tumour diagnosis. Index strokes were due to either symptomatic ischaemic infarction of the brain or symptomatic intracranial haemorrhage. Episodes of transient neurological symptoms without radiological evidence of ischaemic infarction of the brain, such as transient ischaemic attacks, were excluded. Symptomatic intracranial haemorrhage was defined as either intracerebral haemorrhage or subarachnoid haemorrhage or subdural haemorrhage that caused symptoms. Patients were excluded if they had a traumatic subarachnoid haemorrhage or if neuroimaging revealed small-volume post-operative sub-arachnoid or subdural haemorrhage and symptoms were not directly attributed to the haemorrhage. The ICD-10 codes used to identify ischaemic stroke and intracranial haemorrhage were: I60.x, I61.x, I62.x, I63.x and i64x.

Electronic patient notes were reviewed to collect information about: tumour diagnosis, time of stroke, stroke aetiology, investigations and management of stroke, treatment, pre- and post-stroke level of disability measured by the modified Rankin Scale (mRS), number of strokes and date of death. If a patient left the local area, his/her London care records were checked. If the patient left London, then (s)he was lost to follow up. However, the available patient data was included in the analysis.

Stroke recurrence included both clinical presentations of stroke and asymptomatic brain infarction identified on imaging. The time of the asymptomatic brain infarct was calculated as the mid-point between the date of the index stroke and the date when their asymptomatic brain infarction was identified on imaging. Analysis of stroke recurrence excluded patients whose index stroke led directly to their death.

Stroke aetiology for ischaemic strokes were based on the results of the patient’s stroke investigations and neuroimaging reports and followed the Trial of Org 10172 in Acute Stroke Treatment (TOAST) criteria^23^. The aetiology of intracerebral haemorrhage was grouped into intratumoral, hypertensive (deep/infratentorial location and hypertension recorded) and other to reflect the importance of intratumoural haemorrhage in the management of primary intracranial tumours. Haemorrhage was recorded as intratumoural if the neuroradiology report(s) stated this. The aetiology of extratumoural haemorrhage was based on tumour treatment and stroke investigations. Post-operative strokes were defined as strokes occurring within 30 days of surgery. Radiation-induced strokes were defined as patients who had strokes in brain territory supplied by vessels in the radiation field and no other mechanism of stroke identified. Both radiation-induced and post-operative strokes were categorised as either ischaemic or symptomatic intracranial haemorrhage. Strokes that extended or evolved were coded with the initial cerebrovascular event.

### Statistical Analysis

Two-sample data sets with a parametric distribution were analysed with Student’s t-test if the data variances were equal or Welch’s t-test if the data variances were unequal. Non-parametric data were analysed using Wilcoxon rank sum tests. Categorical variables were analysed with Chi-squared testing.

Survival was analysed with Kaplan-Meier methods and log-rank tests, logistic regression (see Supplemental Material for unadjusted odds ratios if the model had more than one explanatory variable) and Cox proportional hazards models. The quality of Cox proportional hazards models was improved by removing explanatory variables using the Akaike Information criterion (see Supplemental Material for initial and final models with results).

Statistical model analysis was performed in R using R statistical software (v4.3.0; R Core Team 2023) on a 64-bit computer running under Windows 11 (64-bit) operating system.

We combined our data with published prevalence data for benign and malignant tumours of the central nervous system to calculate the relative frequency of stroke in patients with benign compared with malignant intracranial tumours in our study. We assumed that the prevalence of benign and malignant primary intracranial tumours in our cohort were the same as in the USA.^10^ The prevalence of benign and malignant primary intracranial tumour in people aged 18 years or older was then calculated by scaling by time the numbers of benign and malignant tumours for the 15 – 39 year age group in the published prevalence data^10^ to an 18 – 39 year age group. The results were then summed with the 40 – 64 and 64+ age groups and the prevalence of benign to malignant primary intracranial tumour calculated from these data.

To estimate the risks of ischaemic stroke and symptomatic intracranial haemorrhage in the six months following diagnosis of a benign intracranial tumour compared with the general population, we assumed that the risks of ischaemic stroke and symptomatic intracranial haemorrhage in the six months following diagnosis of a malignant intracranial tumour in our study cohort were the same as the relative risks reported in a nationwide Swedish study of stroke after diagnosis of a malignant tumour for the same variables.^8^ We calculated the relative risk of ischaemic stroke in the six months following a diagnosis of a benign intracranial tumours compared with the general population as:

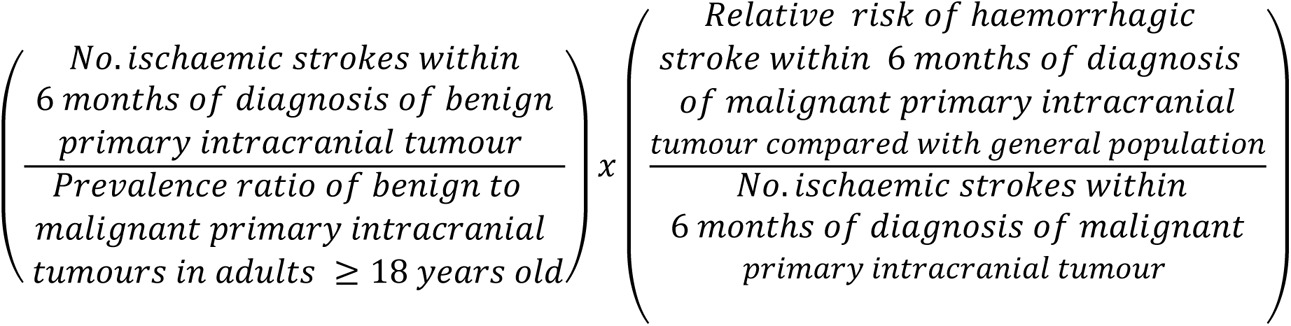

(see Supplemental Material for derivation). The same calculation was made for symptomatic intracranial haemorrhage within 6 months of diagnosis of a benign primary intracranial tumour after replacing “number of ischaemic strokes” with “number of symptomatic intracranial haemorrhages” in the equation.

This study was approved by the King’s College Hospital institutional review board (NEURO_04052022). Anonymised data were used for analysis. Therefore, informed consent was not required. The reporting of this study conforms to the STROBE guidelines (Strengthening the Reporting of Observational Studies in Epidemiology).

## RESULTS

A search of the hospital data base revealed 7854 unique patients who had inpatient stays with an ICD-10 code corresponding to a primary tumour of the brain, cranial nerves or cranial meninges. We identified 333 patients from this cohort who had a clinical diagnosis of stroke within the study period. We excluded patients who had either a stroke before their tumour diagnosis or patients whose clinical diagnosis of stroke was not confirmed on either computer tomography or magnetic resonance imaging. This left 258 patients in our study (Fig. 1, Table 1).

**Figure 1.**
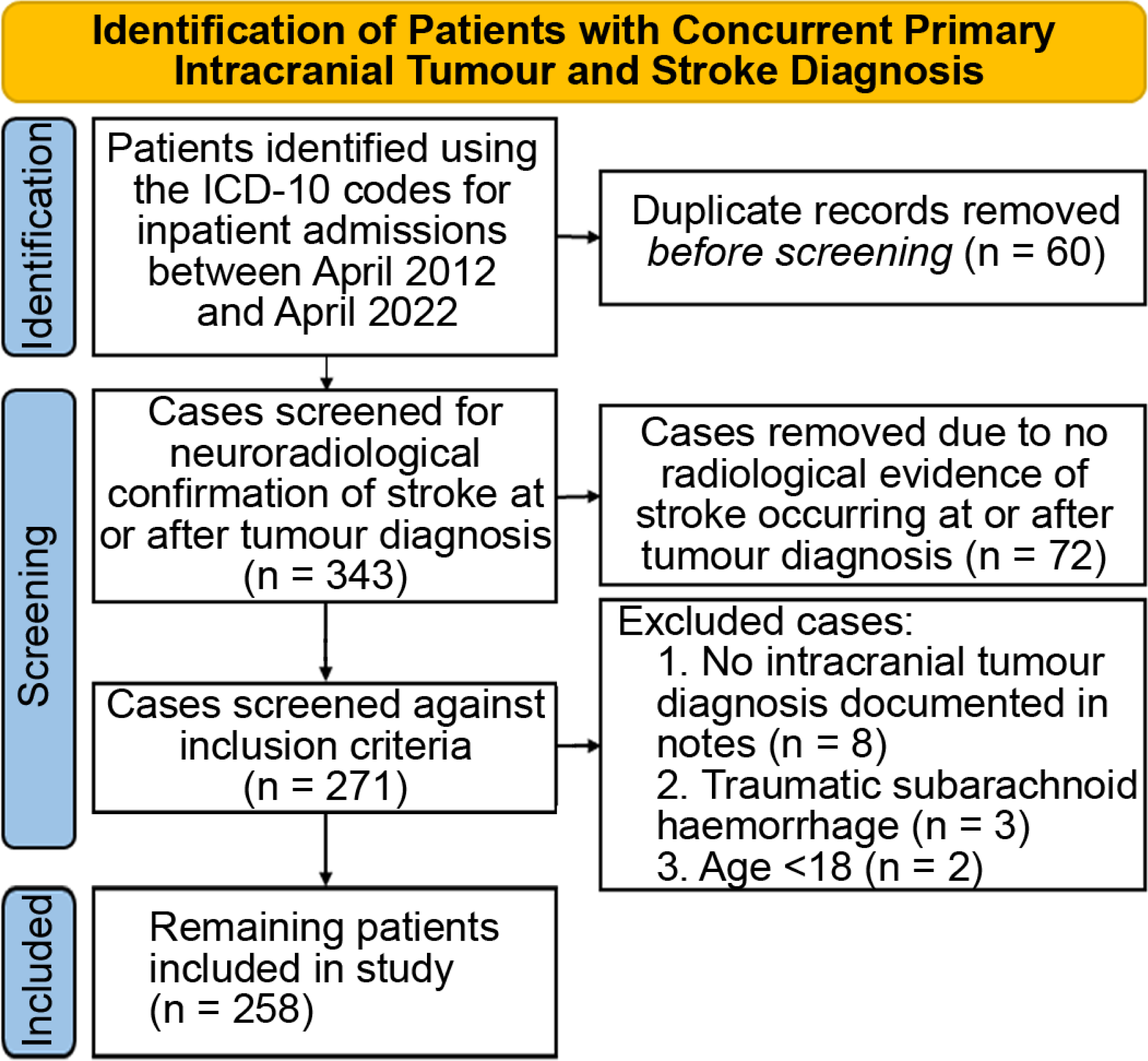
Consort flow diagram. Process to identify patients with primary intracranial tumours who had a stroke at tumour diagnosis or afterwards

**Table 1.**
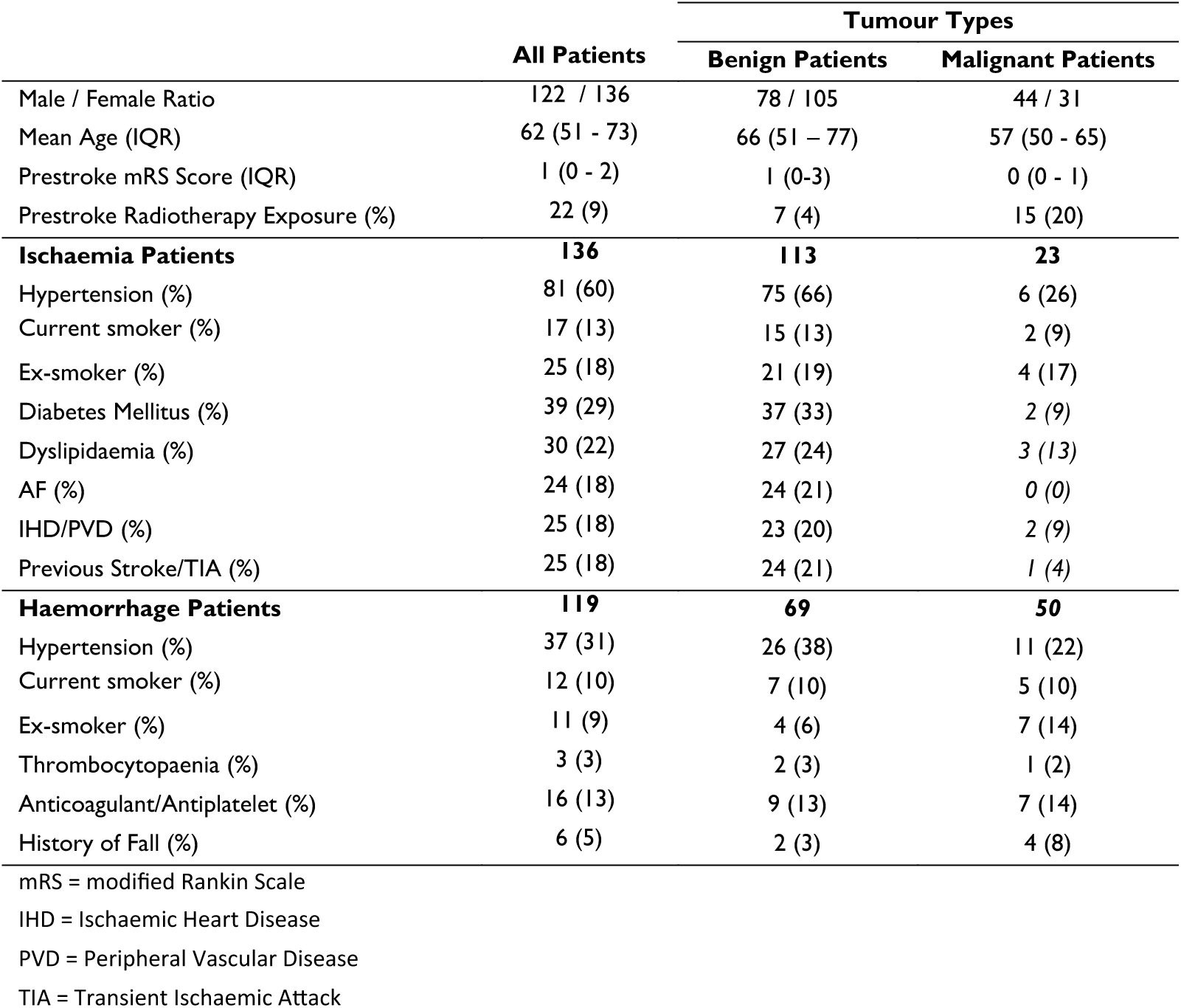
Patient demographics and conventional stroke risk factors

|  | All Patients | Tumour Types |  |
| --- | --- | --- | --- |
|  |  | Benign Patients | Malignant Patients |
| Male / Female Ratio | 122 / 136 | 78 / 105 | 44 / 31 |
| Mean Age (IQR) | 62 (51 - 73) | 66 (51 - 77) | 57 (50 - 65) |
| Prestroke mRS Score (IQR) | 1 (0 - 2) | 1 (0-3) | 0 (0 - 1) |
| Prestroke Radiotherapy Exposure (%) | 22 (9) | 7 (4) | 15 (20) |
| **Ischaemia Patients** | **136** | **113** | **23** |
| Hypertension (%) | 81 (60) | 75 (66) | 6 (26) |
| Current smoker (%) | 17 (13) | 15 (13) | 2 (9) |
| Ex-smoker (%) | 25 (18) | 21 (19) | 4 (17) |
| Diabetes Mellitus (%) | 39 (29) | 37 (33) | 2 (9) |
| Dyslipidaemia (%) | 30 (22) | 27 (24) | 3 (13) |
| AF (%) | 24 (18) | 24 (21) | 0 (0) |
| IHD/PVD (%) | 25 (18) | 23 (20) | 2 (9) |
| Previous Stroke/TIA (%) | 25 (18) | 24 (21) | 1 (4) |
| **Haemorrhage Patients** | **119** | **69** | **50** |
| Hypertension (%) | 37 (31) | 26 (38) | 11 (22) |
| Current smoker (%) | 12 (10) | 7 (10) | 5 (10) |
| Ex-smoker (%) | 11 (9) | 4 (6) | 7 (14) |
| Thrombocytopaenia (%) | 3 (3) | 2 (3) | 1 (2) |
| Anticoagulant/Antiplatelet (%) | 16 (13) | 9 (13) | 7 (14) |
| History of Fall (%) | 6 (5) | 2 (3) | 4 (8) |

mRS = modified Rankin Scale
IHD = Ischaemic Heart Disease
PVD = Peripheral Vascular Disease
TIA = Transient Ischaemic Attack

The index strokes in this cohort comprised 119 (46%) symptomatic intracranial haemorrhages, 136 (53%) ischaemic strokes and 3 (1%) mixed post-operative index strokes, that is a simultaneous infarct and haemorrhage in distinct neurovascular territories identified on neuroimaging following surgery. The median patient age when the index stroke occurred was 62 (IQR 51–73) years (Table 1). Multiple strokes occurred in 68 patients. In total, we identified 337 strokes in 258 patients. This included seven new asymptomatic brain infarctions identified by surveillance imaging of the tumour.

The primary intracranial tumours in our cohort comprised 75 malignant (29% cohort) and 183 benign tumours (71% cohort). The three most common central nervous system tumours were meningioma (42%), glioblastoma (21%) and pituitary adenoma (17%) (Table 2).

**Table 2.**
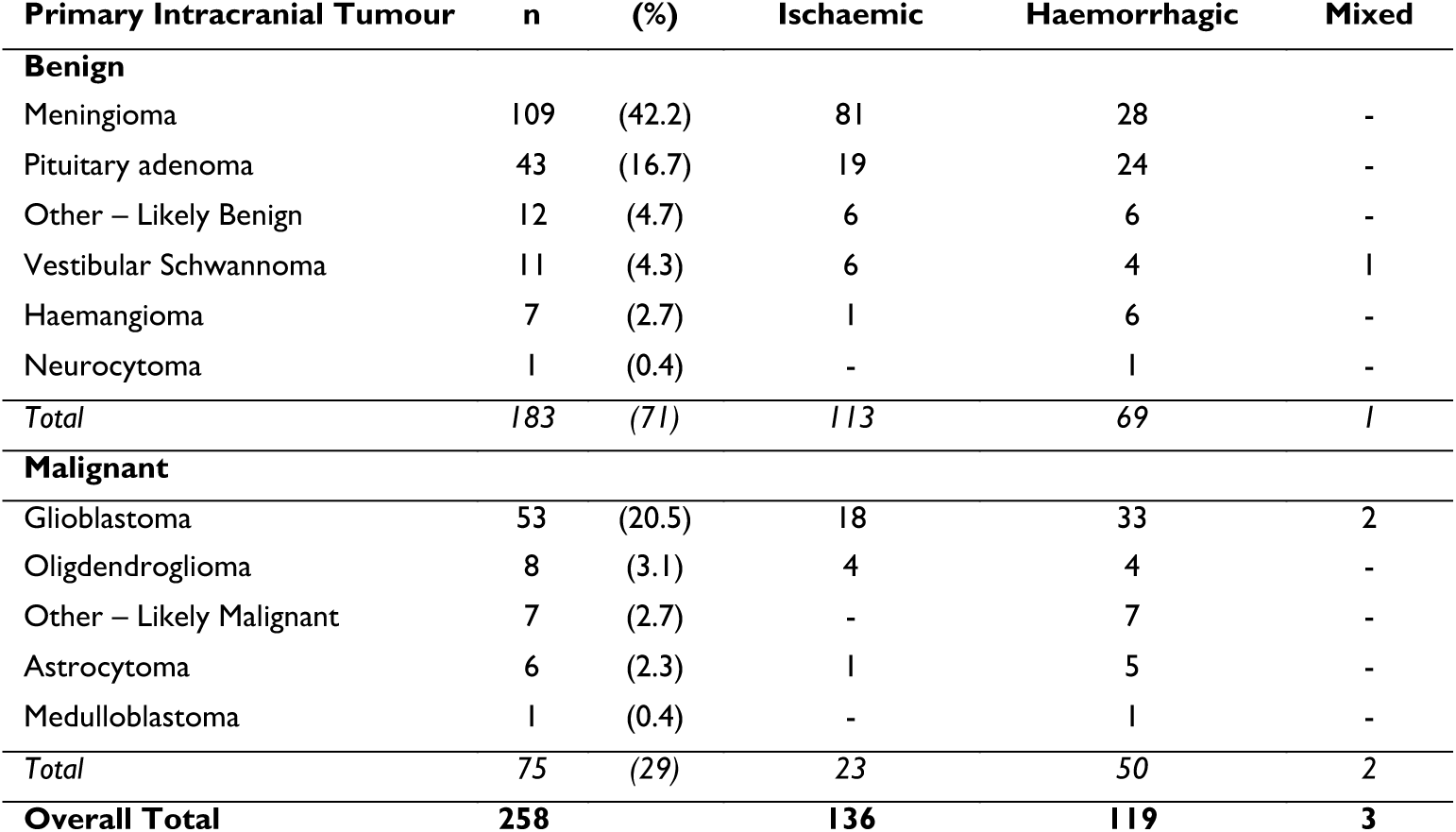
Primary intracranial tumours with stroke types. The tumour categories ‘Other - Likely Benign’ and ‘Other - Likely Malignant’ were used when no tissue diagnosis was recorded in the in the patient’s notes, but treatment for a benign or malignant tumour was documented.

Our data combined with published estimates for the prevalence of benign and malignant primary intracranial tumours^10^ indicated that, overall, patients with malignant primary intracranial tumours were 2.7 times more likely to have a stroke during the study period than patients with benign intracranial tumours (see Methods).

### Stroke type and aetiology

The index stroke in the 75 patients with a malignant primary intracranial tumour comprised 50 symptomatic intracranial haemorrhages, 23 ischaemic strokes and two mixed post-operative strokes (Table 2). In contrast, the index stroke suffered by the 183 patients with a benign primary intracranial tumour consisted of 69 symptomatic intracranial haemorrhages, 113 ischaemic strokes and one mixed post-operative stroke (Table 2). Our data indicated that patients with a malignant tumour were more likely to have a symptomatic intracranial haemorrhage whereas those with a benign tumour were more likely to have an ischaemic stroke (Chi-squared test with Yate’s correction: ꭓ²=18.4, df =1, *P*=1.8x10⁻^5^).

We explored the reasons for the high rate of symptomatic intracranial haemorrhage in patients with intracranial tumours. The majority (84/119, 71%) of haemorrhages were either intratumoural (n=46) or attributable to tumour treatment (31 post-operative and 7 radiation-induced)(Table 3). Only 6/119 (5%) haemorrhages were attributed to hypertension.

**Table 3.** Aetiology of ischaemic and symptomatic intracranial haemorrhages in patients with a primary intracranial tumour.

|  | n | (%) |
| --- | --- | --- |
| **Haemorrhagic** |  |  |
| Intratumoural | 46 | (38.7) |
| Post-operative | 31 | (26) |
| Sub-arachnoid | 10 | (8.4) |
| Radiation-induced | 7 | (5.9) |
| Hypertensive | 6 | (5) |
| Other | 19 | (16) |
| **Total** | **119** | **(100)** |
| **Ischaemic** |  |  |
| Post-operative | 51 | (37.5) |
| Cardioembolic | 28 | (20.6) |
| Small vessel disease | 23 | (16.9) |
| Undetermined | 21 | (15.4) |
| Radiation Induced | 6 | (4.4) |
| Other | 5 | (3.7) |
| Large Vessel Atherosclerosis | 2 | (1.5) |
| **Total** | **136** | **(100)** |

*Note - the three patients with mixed strokes were not included in the above table*

The most common aetiology for ischaemic strokes was post-operative (51/136, 38%) (Table 3). The most common non-surgical causes were cardioembolic (28/136, 21%) and small vessel disease (23/136, 17%). Ischaemic strokes were more likely to occur in the same vascular territory as the tumour when the patient had a malignant tumour compared with a benign tumour (malignant 15/23 ischaemic strokes, benign 25/112 ischaemic strokes; Chi-squared test with Yate’s correction, ꭓ²=15.1, df=1, *P*=0.001).

Radiotherapy is associated with an increased risk of stroke^12,19,24^ and could contribute to patients having a stroke in the same vascular territory as the tumour. In our cohort, 54/258 (21%) patients received radiotherapy as part of their tumour treatment. 22/54 (41%) radiotherapy patients had their initial stroke after their radiotherapy had finished. Nine strokes were attributed to other aetiologies, leaving 13 strokes (seven haemorrhagic, six ischaemic) that fulfilled our criteria for radiation-induced stroke (see Methods).

### Time of index stroke

We investigated when the index stroke occurred with respect to tumour diagnosis (Fig. 2A). The median time from tumour diagnosis to index stroke was shorter for symptomatic intracranial haemorrhage than ischaemic strokes (haemorrhagic 26 (IQR 0–210) days, ischaemic 139 (IQR 2–1275) days; Wilcoxon rank sum test, *P* = 4x10^-4^)(Fig. 2B).

**Figure 2.**
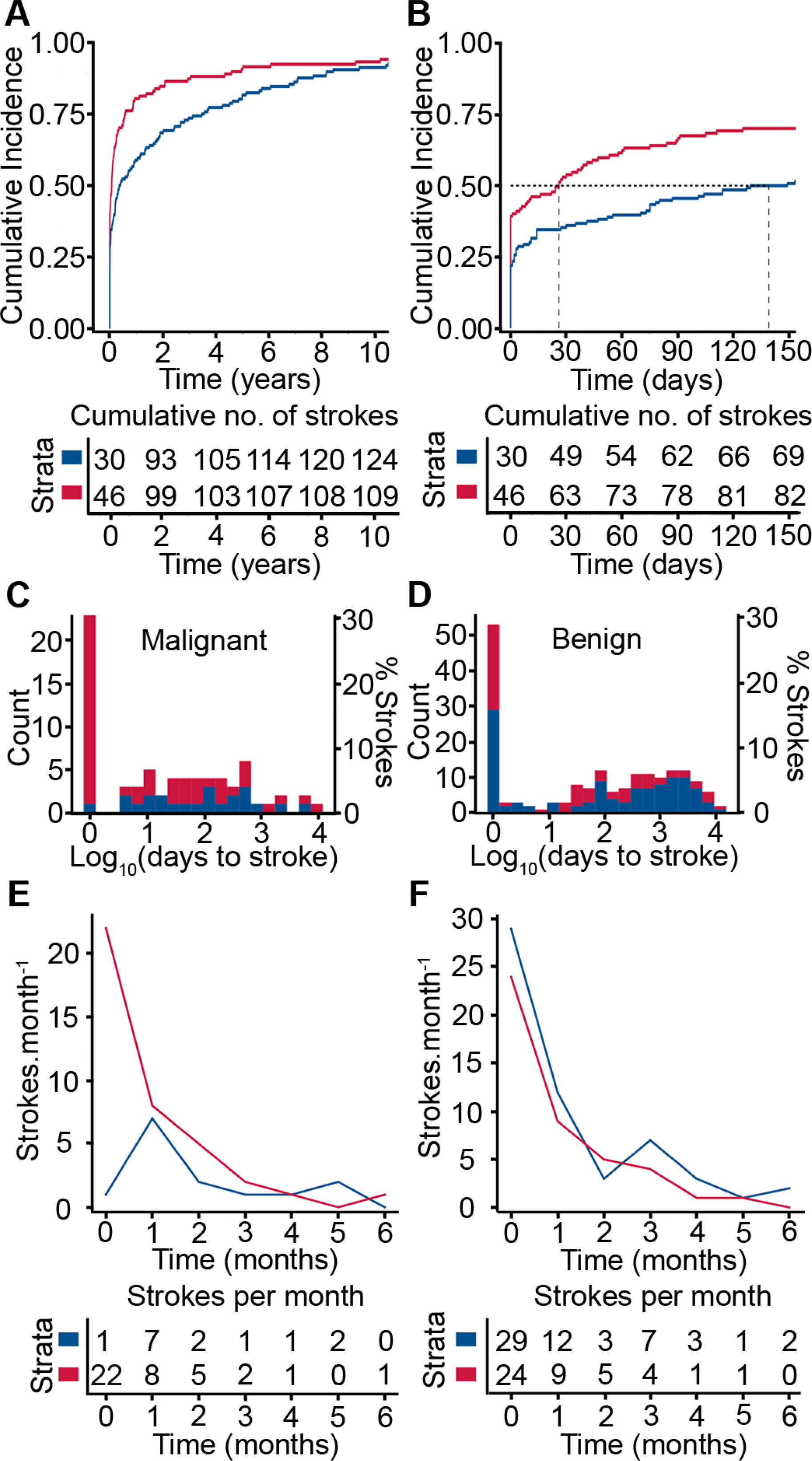
Time of index stroke after tumour diagnosis. **(A)** Cumulative incidence of all haemorrhagic (red) and all ischaemic (blue) stroke following intracranial tumour diagnosis over the 10-year study period. **(B)** Cumulative incidence of all haemorrhagic (red) and all ischaemic (blue) strokes in the 150 days following tumour diagnosis. Vertical dashed lines denote median times from diagnosis to symptomatic intracranial haemorrhage and ischaemic stroke. **(C)** Histogram of times of ischaemic (blue) and haemorrhagic (red) strokes after diagnosis of malignant primary intracranial tumour. **(D)** Histogram of times of ischaemic (blue) and haemorrhagic (red) strokes after diagnosis of benign primary intracranial tumour. Note log scale on x-axis in **C** and **D**. **(E)** Malignant primary intracranial tumours, ischaemic strokes (blue) and symptomatic intracranial haemorrhage (red) at tumour diagnosis (time 0) and monthly stroke rates after tumour diagnosis. **(F)** Benign primary intracranial tumours, ischaemic strokes (blue) and symptomatic intracranial haemorrhage (red) at tumour diagnosis (time 0) and monthly stroke rates after tumour diagnosis.

Stroke occurred at tumour diagnosis with equal frequency for malignant and benign primary intracranial tumours (malignant 23/75 (31%) patients; benign 53/183 (29%) patients; ꭓ^2^=0.035, df=1, *P*=0.85) (Fig. 2C, D). The most common primary brain tumours associated with a symptomatic intracranial haemorrhage index stroke were glioblastoma (16/33, 48%), pituitary adenoma (13/24, 54%) and meningiomas (28/109, 26%) (Table 2). Overall, patients with a malignant primary intracranial tumour were more likely to have symptomatic intracranial haemorrhage (22/23, 96%) at tumour diagnosis than patients with a benign tumour (24/53, 45%) (ꭓ^2^=15.0, df=1, *P*=1x10^-4^)(Fig. 2C, D).

One third of all strokes (82/258) occurred in the 30-day post-operative period. The median time from tumour surgery to the index stroke in the postoperative period was 2 (IQR 1–6) days.

Epidemiological studies of stroke in patients with cancer including malignant primary intracranial tumours have revealed a spike in incidence in the months around cancer diagnosis.^8,15^ We found that the incidence of symptomatic intracranial haemorrhage was highest at tumour diagnosis and decreased in the following months in patients with malignant primary intracranial tumours (Fig. 2E). Ischaemic stroke was most common in the month following diagnosis of a malignant primary intracranial tumour and then decreased (Fig. 2E)

Analysis of stroke rates in the period following diagnosis of a benign intracranial tumour also showed a spike in incidence at tumour diagnosis and in the following months for both ischaemic stroke and symptomatic intracranial haemorrhage (Fig. 2F). We estimated that the risk of ischaemic stroke and symptomatic intracranial haemorrhage in the six months at or after diagnosis of a benign primary intracranial were respectively 2.5 and 5.0 times that of the general population (see Methods). Strokes at the diagnosis of benign tumours may be coincidental, with the tumour being detected as a consequence of neuroimaging investigations for stroke. Therefore, we repeated the risk calculation after excluding all the ischaemic strokes that occurred at tumour diagnosis. This gave a lower bound of 1.3 to the risk of ischaemic stroke in the six months at or after diagnosis of a benign intracranial tumour.

### Stroke-associated mortality

We explored survival of patients with a primary intracranial tumour following a stroke. The median overall survival after stroke for all patients with a malignant intracranial tumour (0.7 [IQR 0.2–1.7] years) was shorter than patients with a benign tumour (1.6 [IQR 0.3–3.8] years; log-rank test, *P*=2.7x10^-8^) (Fig. 3A). However, the survival curves for patients with benign and malignant primary intracranial tumours overlap for the 1-2 months following hospital admission and then diverge (Fig. 3B). This suggested that early mortality following a stroke was dominated by the effect of the stroke. Accordingly, we analysed mortality in more detail by dividing patients into those that died in hospital following admission without being discharged and patients who were discharged following their stroke.

**Figure 3.**
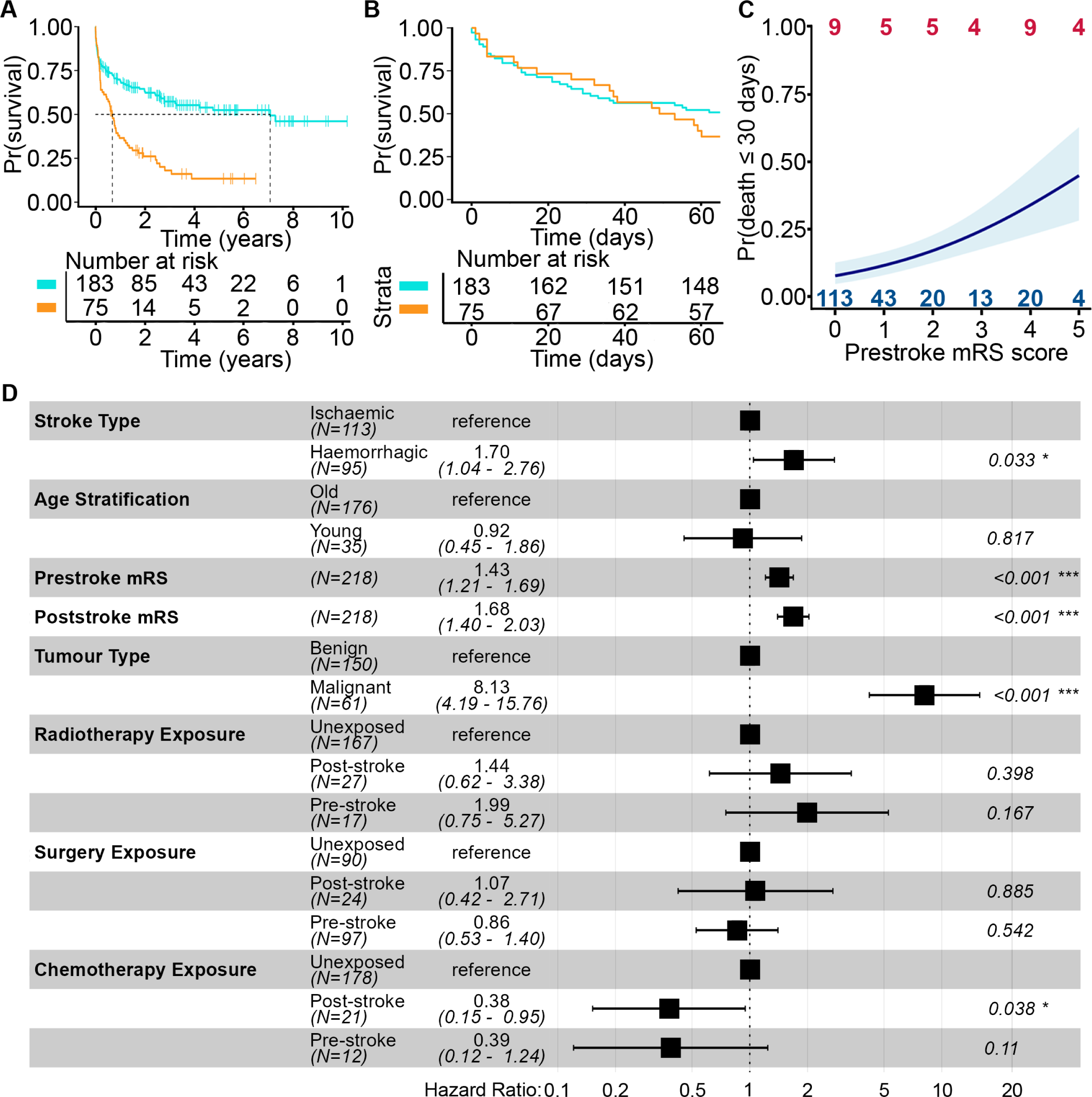
Death following stroke in patients with primary intracranial tumours. **(A)** Kaplan-Meier plot of survival following stroke for all patients with a malignant (orange) or benign (light blue) primary intracranial tumour. Dashed lines indicate median survival times. (**B**) Kaplan-Meier plot of 60-day survival following stroke for patients with a malignant (orange) or benign (light blue) tumour. (**C**) Probability of death within 30 days of stroke estimated from logistic regression model. Light blue shading denotes the standard error of the logistic regression plot. Blue numbers (top), patients who survived 30 days after their stroke; red numbers (bottom), patients who died within 30 days of their stroke for each prestroke mRS score. (**D**) Forest plot of hazard ratios for explanatory variables in final Cox Proportional Hazards model of survival after hospital discharge.

A total of 40/258 (16%) patients died in hospital following their first stroke. The median survival for patients who died in hospital was 12 (IQR 4–25) days. There was no difference in inpatient mortality rates between patients with benign or malignant tumours (benign 28/178 patients, malignant 12/72 patients; ꭓ^2^=0.03, df=1, *P*=0.86)(Fig. 3B) and no difference in inpatient survival duration (Wilcoxon rank sum test, *P*=0.84).

We quantified early mortality by using death within 30 days of the index stroke (benign, 28 patients; malignant, 9 patients). Logistic regression revealed that the patient’s level of disability before the stroke was associated with increased early death after stroke (Adjusted OR per unit increase in mRS score 1.47 [95% CI, 1.16-1.88]) (Fig. 3C). Age at the time of stroke (Adjusted OR per year 1.03 [95% CI, 1.00-1.05]) had a modest effect. We found no evidence that malignancy of the tumour (malignant/benign Adjusted OR 1.12 [95% CI, 0.43-2.80]) or stroke type had an effect (ischaemic/haemorrhagic Adjusted OR 0.53 [95% CI, 0.22-1.21])(Table S1).

Next, we analysed the survival of the 218/258 (84%) patients who were discharged from hospital following their stroke (Fig. S1A). We used the level of disability at hospital discharge, which is a composite measure of stroke severity and stroke recovery, to analyse outcomes for discharged patients. We found that survival of discharged patients was affected by: the level of disability before the stroke (hazard ratio (HR) for death per unit increase in mRS score = 1.43 [95% CI, 1.21–1.69]); the level of disability at hospital discharge (HR per unit increase in mRS score = 1.68 [95% CI, 1.40–2.03]); whether the stroke was haemorrhagic rather than ischaemic (HR=1.70 [95% CI, 1.04–2.76]); and whether the tumour was malignant (HR=8.13 [95% CI, 4.19–15.76])(Figs. 3D, S1B).

### Stroke-induced morbidity

We next explored the effect of stroke on the level of disability of patients who were discharged from hospital. The level of disability increased markedly following a stroke. On average, patients with a malignant tumour had better function than patients with benign tumours before their stroke (median mRS score: malignant=0, benign=1; Wilcoxon Rank sum test, *P*=0.003; Fig. 4A), but had similar levels of disability after their stroke (median mRS score: malignant=4, benign=4; Wilcoxon Rank test, *P*=0.53) (Fig. 4B). Only 37% of patients with a malignant tumour who were independent before their stroke remained independent at hospital discharge. The proportion of patients who were independent before their stroke and became dependent after their stroke was higher in the malignant tumour group than the benign tumour group (malignant 45/71 (63%), benign 80/167 (48%), χ^2^=5.99, df=1, *P*=0.014).

**Figure 4.**
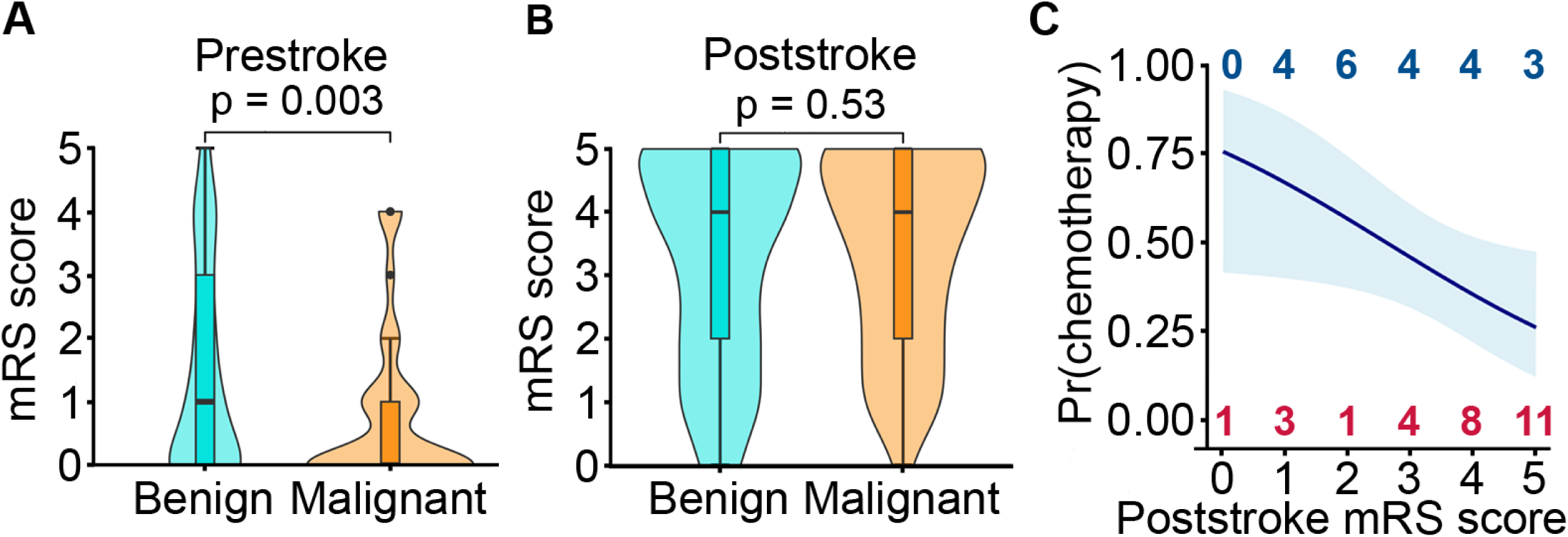
Stroke-induced disability and consequences. **(A)** Violin plot of level of disability before stroke in patients with benign (light blue) and malignant intracranial tumours (orange). **(B)** Violin plot of level of disability after stroke in patients with benign (light blue) and malignant intracranial tumours (orange) who were discharged from hospital. **(C)** Probability of patients with a malignant intracranial tumour receiving chemotherapy estimated from logistic regression model. Light blue shading denotes the standard error of the logistic regression plot. For each post-stroke mRS value: blue numbers (top), patients who received chemotherapy after their stroke; red numbers (bottom), patients that did not receive chemotherapy.

A patient’s suitability for cancer treatment is negatively impacted by disability. Therefore, we investigated if a stroke affected whether patients with a malignant primary intracranial tumour subsequently had a first course of chemotherapy. We found that there was an inverse relationship between probability of receiving chemotherapy and the patient’s post-stroke mRS score (OR per unit increase in mRS 0.65 [95% CI, 0.42-0.96]) (Fig. 4C).

### Stroke recurrence

The median time from index stroke to stroke recurrence was 64 (IQR 3-325) days (Figs. 5A-B, S2A). The cumulative incidences of a second clinically-symptomatic stroke at 1 month, 3 months, 6 months and 1 year were 12%, 14%, 17% and 22% respectively (Fig. S2B). When asymptomatic brain infarction was included, the cumulative incidences of a second stroke at 1 month, 3 months, 6 months and 1 year were 14%, 17%, 21% and 23% respectively (Fig. 5B).

**Figure 5.**
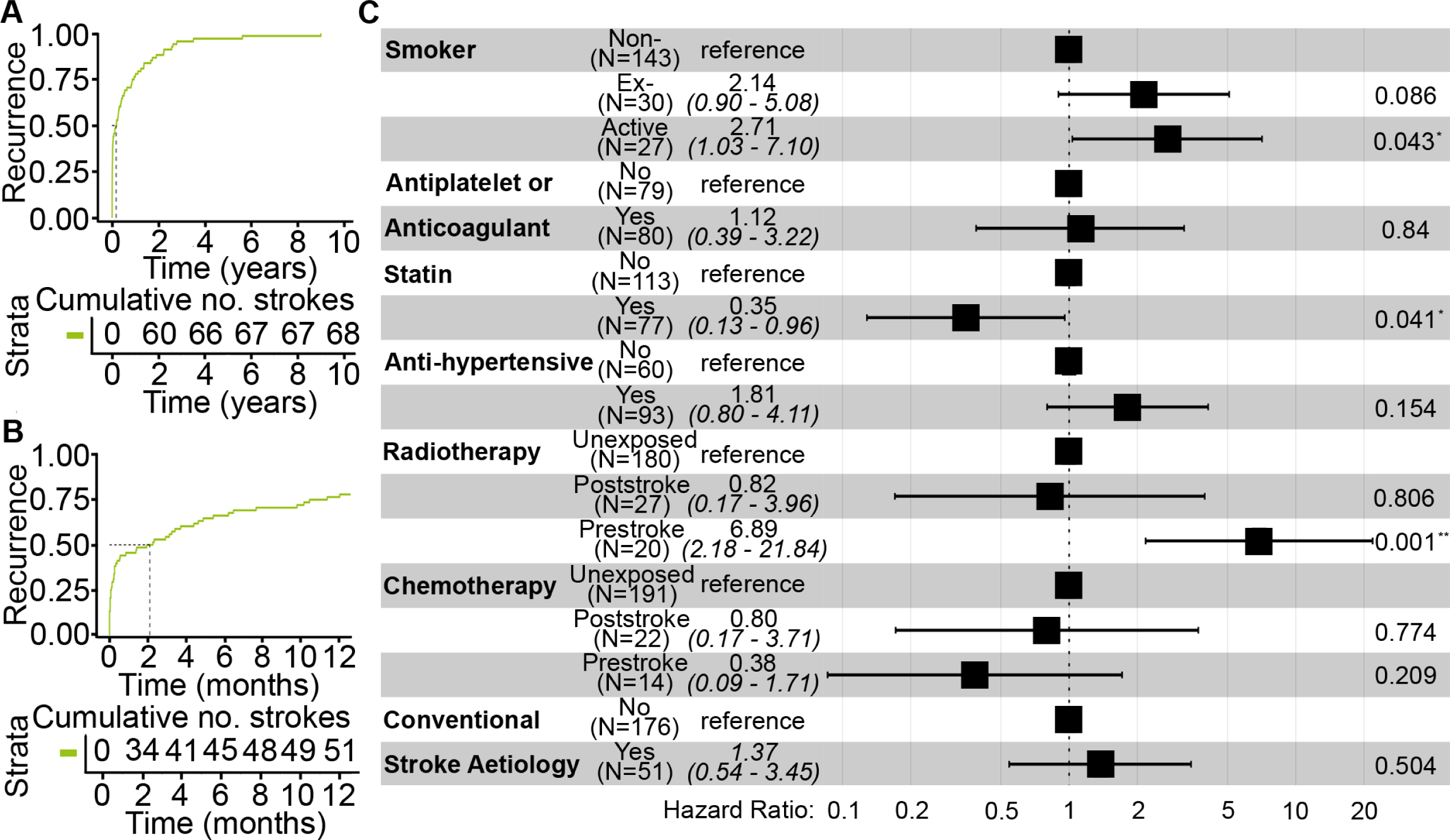
Recurrent strokes. **(A)** Cumulative incidence plot of patients having a recurrent stroke including asymptomatic brain infarction over the study period. **(B)** Cumulative incidence plot of patients having a recurrent stroke including asymptomatic brain infarction in the first year after their index stroke. **(C)** Forest plot of hazard ratios of explanatory variables in statistical model of stroke recurrence.

Ischaemic second strokes were more common than haemorrhagic second strokes (ischaemic 53 patients, haemorrhagic 15 patients; binomial test, *P*=2x10^-5^). Recurrent asymptomatic brain infarction was identified in seven patients on tumour surveillance imaging. The maximum number of strokes an individual patient had during the study period was four.

We next considered the relationship between the index and second strokes. The 30 patients who had a haemorrhagic index stroke subsequently had 11 haemorrhagic and 19 ischaemic second strokes. The 38 patients who had an ischaemic index stroke subsequently had 4 haemorrhagic and 34 ischaemic strokes. We concluded that patients who had an ischaemic index stroke were more likely to have an ischaemic recurrent stroke (ꭓ^2^=5.23, df=1, *P*=0.022).

We investigated the stroke-related and tumour-related risk factors for stroke recurrence. Our analysis included an explanatory variable that grouped all patients that had a stroke aetiology conforming to the TOAST criteria or hypertension to explore whether conventional stroke aetiologies were associated with increased the risk of stroke recurrence in patients with primary intracranial tumours. We found that the risk of recurrent strokes was higher in patients who had radiotherapy prior to their first stroke (HR=6.59 [95% CI, 2.09-20.8]) or who were an active smoker (HR=2.86 [95% CI, 1.09-7.5])(Fig. 5C), but was not affected by whether: the index stroke was ischaemic or symptomatic intracranial haemorrhage (ischaemic 38 patients, haemorrhagic 30 patients; HR=1.05 [95% CI, 0.38-2.9]); the tumour was malignant (HR=0.60 [95% CI, 0.15-2.4]) or the stroke was attributed to a conventional stroke aetiology (HR=1.4 [95% CI, 0.4-3.5])(Fig. S2A).

Receiving a statin after the index stroke was associated with a lower risk of recurrent stroke (HR=0.36 [95% CI, 0.13-0.98]) (Figs. 5C, S2A).

### Stroke treatment

Eleven patients (10 benign tumours, 1 undiagnosed malignant tumour) underwent thrombolysis for ischaemic stroke. 51 (43%) patients with intracerebral haemorrhage and three patients with an ischaemic stroke causing mass effect required surgery.

We assessed stroke risk factor optimisation in our cohort (Table 1). We found no difference in the mean arterial blood pressure of patients with benign or malignant tumours and symptomatic intracranial haemorrhage (mean arterial pressure mm Hg (standard deviation): benign 97 (10), malignant 95 (13); t-test, *P*=0.47, n=60 haemorrhages). Screening blood tests, such as HbA1c and lipid profile, were performed in under 50% of patients during their inpatient admission (Table S3).

We investigated secondary prevention of ischaemic stroke after excluding patients who had haemorrhagic transformation. 76/124 (61%) patients received a statin and 86/124 (69%) patients were prescribed either an antiplatelet agent or anticoagulant. 25/43 (58%) patients with recurrent ischaemic strokes were started on an antiplatelet agent prior to their second stroke.

## DISCUSSION

We conducted a retrospective cohort study of stroke in patients with primary intracranial tumours who attended a centre with a regional Neuro-oncology unit and a regional stroke unit. Our patient cohort captured all stroke types and severities in addition to being representative of the primary intracranial tumours prevalent in the general population.^25^ Our findings suggested that stroke is a wider issue for patients with primary intracranial tumours than is commonly recognised. The incidence of stroke has been reported to be elevated in the months following diagnosis of malignant primary intracranial tumours.^8,15^ Our data indicated that the risk of stroke was also raised in the six months following diagnosis of benign intracranial tumours. Stroke was a major cause of mortality and neurological disability in people with benign or malignant primary intracranial tumours. The 30-day mortality following stroke and stroke recurrence rates were equally high for benign and malignant intracranial tumours. Taking a statin after the index stroke was associated with lower stroke recurrence. Notably, stroke had consequences for tumour treatment: the marked increase in disability caused by stroke was associated with a reduced likelihood that patients with malignant intracranial tumours would receive chemotherapy.

The malignancy of the tumour affected stroke type. Symptomatic intracranial haemorrhage was more commonly associated with malignant primary intracranial tumours. The haemorrhages were predominantly intratumoural or related to tumour treatment and were not clearly attributable to hypertension as in the general population.^26^ The precise mechanism for the intratumoural haemorrhage is debated, but likely relates to neovascularization in malignant tumours.^27^

The in-hospital mortality following stroke in patients with primary intracranial tumours was similar to the all-cause, in-hospital mortality following stroke reported for the general population.^28^ The level of disability before stroke and the severity of the stroke^29,30^ were major determinants of 30-day mortality and of death after hospital discharge. Having a malignant tumour was not a risk factor for 30-day death. We concluded that initial management of the stroke should focus on stroke treatment to improve survival while taking account of the patient’s intracranial tumour, for example planning the start and stop times of anti-platelet therapy around brain tumour surgery.

There was a marked increase in disability in all patients who were discharged from hospital. The increase in disability was associated with a lower likelihood that patients with malignant intracranial tumours would receive chemotherapy. Hence, stroke in patients with malignant primary intracranial tumours may shorten patients’ lives directly and, also, by precluding tumour treatment.

We estimated that the risk of stroke in the six months at or after diagnosis of a benign intracranial tumour was 1.3 – 2.5 times the general population. Several factors may affect the accuracy of our estimates. Prevalence data for benign and malignant tumours had to be calculated from incidence data and survival curves.^10^ If benign intracranial tumours were underreported, then we would have overestimated stroke risk. Differences in the demographics of our study population (UK) and the populations that were the basis for the prevalence data (USA)^10^ and risk of stroke after tumour diagnosis (Sweden)^8^ may have introduced further error into our results.

The spike in incidence of ischaemic strokes and symptomatic intracranial haemorrhage following diagnosis of a benign intracranial tumour mirrored the time course of the increased incidence of stroke in the months after the diagnosis of many malignant tumours, including malignant primary tumours of the central nervous system.^15,31^ The reasons are unclear. Brain scans arranged as a stroke investigation would also detect a “co-incidental” benign intracranial tumour. However, coincidence does not explain the time course of the increase in stroke risk in the months after tumour diagnosis. The tumour or its treatment may be explanatory factors. Benign intracranial tumours can affect the brain, for example by causing intracerebral oedema or epileptic seizures.^3^ Hence, it remains possible that benign intracranial tumours increase the risk of stroke through as yet undefined mechanisms. This is supported by the high rates of stroke recurrence in our patients with benign intracranial tumours.^28,32,33^ Investigating this further will require studying stroke in distinct types of benign intracranial tumour.

The treatments for primary intracranial tumours, particularly surgery and radiotherapy, are risk factors for stroke. The frequency of post-operative strokes that we report is less than in other studies^12,13^, possibly because of enhanced neurosurgical efforts to reduce surgery-related stroke. We found fewer radiotherapy-induced strokes than reported in other studies.^12,13^ However, we are likely to have underestimated radiotherapy-induced strokes because our definition of a radiation-induced stroke was strict, requiring that no other cause for the stroke was found.

Ischaemic strokes were more likely to occur in the vascular territory of malignant tumours compared with benign tumours. Multiple factors may contribute to this finding including: compression of blood vessels; procoagulant effects of cancer and radiotherapy; and higher metabolic demands of malignant tumour.^12,13,24,34,35^

Stroke recurrence rates for patients with benign intracranial tumours and patients with malignant primary intracranial tumours were similar to stroke recurrence rates in patients with active cancer originating outside the brain^36^. These rates are high compared with the general population.^28,32,33^ Hence, our data suggest that patients with benign primary intracranial tumours have a high stroke recurrence rate. Radiotherapy treatment and active smoking are risk factors for recurrent stroke in patients with primary intracranial tumours as described for the general population.^26^ However, it is not clear that these factors account for all of the increase in stroke recurrence. A larger study looking at stroke recurrence in distinct types of primary intracranial tumour is needed to investigate this issue.

Stroke prevention is vital not only for maximizing quality of life, but also to ensure patients’ performance status remains above the threshold required for tumour treatment. Standard stroke risk factors should be optimized. In addition to stopping smoking, our data suggest that taking a statin after a stroke was associated with a lower risk of recurrent stroke. This is consistent with a protective effect of statins against stroke, which has been reported for patients who have radiotherapy for head and neck cancer.^37,38^ The mechanism of the protective effect of statins remains to be elucidated.

Ischaemic index strokes were more likely to be followed by recurrent ischaemic stroke rather than by symptomatic intracranial haemorrhage. Hence, an antiplatelet agent should be considered after an ischaemic index stroke, particularly if the patient has had radiotherapy, despite the high risk of symptomatic intracranial haemorrhage.

The number of patients receiving secondary stroke prevention treatment was lower than expected. Anti-stroke prophylaxis may be deferred whilst patients undergo tumour treatment, such as surgery. This factor combined with multiple teams providing clinical care may result in anti-stroke prophylaxis not being initiated or restarted. Education of non-stroke clinicians caring for patients with primary intracranial tumours would help to prevent this.

Management of stroke in patients with a primary intracranial tumour is challenging because stroke treatments may cause tumour-related side-effects, such as haemorrhage. These patients would benefit greatly from greater attentions to stroke management and stroke prevention as this will reduce their disability, optimise their quality of life and ensure that tumour treatment options remain open.

## Data Availability

Anonymised data will be made available upon reasonable request to the Authors

## ACKNOWLEDGEMENTS

For the purposes of open access, the author has applied a Creative Commons Attribution (CC BY) licence to any Accepted Author Manuscript version arising from this submission. We thank Dr Jonathan Best and Dr Yee-Mah Haur for constructive comments.

## SUPPLEMENTAL MATERIAL

STROBE checklist, Supplemental Methods, Figures S1-S2,

Tables S1-S3.

## Non-standard Abbreviations and Acronyms

mRS: modified Rankin Scale
TOAST: Trial of Org 10172 in Acute Stroke Treatment

